# A tipping point in cancer-immune dynamics leads to divergent immunotherapy responses and hampers biomarker discovery

**DOI:** 10.1101/2020.10.29.20222455

**Authors:** Jeroen H.A. Creemers, W. Joost Lesterhuis, Niven Mehra, Winald R. Gerritsen, Carl G. Figdor, I. Jolanda M. de Vries, Johannes Textor

**Affiliations:** Department of Tumor Immunology, Radboud Institute for Molecular Life Sciences; Radboudumc, Nijmegen, The Netherlands; Department of Medical Oncology, Radboudumc, Nijmegen, The Netherlands; School of Biomedical Sciences and Telethon Kids Institute, University of Western Australia, Perth, Australia; Oncode Institute, Nijmegen, The Netherlands; Data Science Department, Institute for Computing and Information Sciences, Radboud University, Nijmegen, The Netherlands

**Author notes:** Corresponding author: Dr. J. Textor, Department of Tumor Immunology, Radboud Institute for Molecular Life Sciences, Radboudumc, Geert Grooteplein 26, 6500 HB Nijmegen (P.O. Box 9101), The Netherlands.

## Abstract

**Background:** Predicting treatment response or survival of cancer patients remains challenging in immuno-oncology. Efforts to overcome these challenges focus, among others, on the discovery of new biomarkers. Despite advances in cellular and molecular approaches, only a limited number of candidate biomarkers eventually enter clinical practice.

**Methods:** A computational modeling approach based on ordinary differential equations was used to simulate the fundamental mechanisms that dictate tumor-immune dynamics and to investigate its implications on responses to immune checkpoint inhibition (ICI) and patient survival. Using *in silico* biomarker discovery trials, we revealed fundamental principles that explain the diverging success rates of biomarker discovery programs.

**Results:** Our model shows that a tipping point – a sharp state transition between immune control and immune evasion – induces a strongly non-linear relationship between patient survival and both immunological and tumor-related parameters. In patients close to the tipping point, ICI therapy may lead to long-lasting survival benefits, whereas patients far from the tipping point may fail to benefit from these potent treatments.

**Conclusion:** These findings have two important implications for clinical oncology. First, the apparent conundrum that ICI induces substantial benefits in some patients yet completely fails in others could be, to a large extent, explained by the presence of a tipping point. Second, predictive biomarkers for immunotherapy should ideally combine both immunological and tumor-related markers, as a patient’s distance from the tipping point can typically not be reliably determined from solely one of these. The notion of a tipping point in cancer-immune dynamics helps to devise more accurate strategies to select appropriate treatments for cancer patients.

## INTRODUCTION

Immunotherapies are revolutionizing clinical care for cancer patients. The most widely used approach, immune checkpoint inhibition (ICI), can lead to long-term survival benefits in patients with advanced melanoma (1), lung cancer (2), and renal cell carcinoma (3). However, not all patients benefit from ICI therapy, and adequate predictions of treatment response have proven elusive so far (4, 5). Efforts to improve these predictions focus mainly on discovering biomarkers in aberrant molecular pathways within the tumor microenvironment that drive immunosuppression and therapeutic resistance (6, 7). These include genomic alterations in oncogenic drivers, the absence of tumor-specific antigens, and the presence of immunosuppressive molecules or cells (8, 9). Despite substantial efforts, only a limited fraction (according to one estimate, <1% (10)) of proposed cancer biomarkers find their way into the clinical practice. These apparent challenges in identifying biomarkers for immunotherapy and translating them into clinical practice could be a consequence of the inherent complexity of cancers and their interaction with the immune system.

To unravel the complexities of cancers and their treatments, researchers have adopted mathematical and computational approaches to complement laboratory research. A plethora of modeling approaches are available, ranging from simple one-variable equations to complex spatial agent-based simulation models. *In silico* modeling has contributed to fundamental insights into tumor growth and cancer progression (11-13), tumor-immune control (e.g., neoantigen prediction as targets for immunotherapy) (14), identification of tumor-associated genes (15), verification of treatment-related safety concerns such as hematological toxicity (16), prediction of treatment responses to chemo- and immunotherapy (17-19), investigation of drug-induced resistance (20), and timing of anti-cancer treatments (21-23). In the context of disease course dynamics, ordinary differential equation (ODE) models have proven useful over the years. ODE models follow the principle that a model should be “as simple as possible but not simpler”. Based on plausible biological assumptions, they aim to reduce the complex reality of the modeled system to its bare essentials to enable the investigation of critical underlying dynamics. The field of quantitative systems pharmacology is built upon this premise. Classically, experimentally-derived pharmacokinetic and pharmacodynamic parameters serve as input for ODE models to investigate the emergent properties of biological systems and to study its consequences in terms of clinical outcomes (24). As an illustrative example, Fassoni *et al*. used ODE models to predict that dose de-escalation of tyrosine kinase inhibitors targeting the oncogenic protein BCR-ABL1 in patients with chronic myeloid leukemia (chronic phase) does not lead to worse long-term outcomes (25). The recent results of the DESTINY trial support this prediction (26).

In this study, we investigate the consequences of tumor-immune dynamics on patients’ responses to ICI and survival in an ODE model. Our model reveals a tipping point within tumor-immune dynamics – a critical threshold for survival culminating in an all-or-nothing principle – that has profound implications for a patient’s disease course and outcome. We show how the presence of a tipping point alone robustly induces heterogeneous immunotherapy treatment outcomes, and how this complicates the search for both prognostic and predictive biomarkers.

## METHODS

### Capturing core mechanisms of tumor development in a mathematical model

We constructed a mathematical model consisting of a system of ordinary differential equations (ODE) to capture essential interactions between cancer cells and lymphocytes during tumor formation. Our model represents tumorigenesis in patients, starting with the malignant transformation of a single cell. The model consists of five equations that describe essential processes in the tumor microenvironment and the lymphatic organs (Figure 1A). In the tumor microenvironment, tumor growth (Equation 1^a^) and T cell-mediated killing of tumor cells (Equation 1^b^) determine the evolution of the tumor burden (the numbers of the equations correspond to those used in Figure 1A). Tumor-infiltrating lymphocytes migrate from the lymph nodes to the microenvironment (Equation 2). Before migration, T cells expand clonally in the lymph nodes’ T cell zones (Equation 3) after conversion of naive T cells into antigen-specific effector T cells (Equation 4). Below, we provide in-depth descriptions of each model equation. We modeled tumor growth – i.e., the formation of tumor cells during carcinogenesis – with the generalized exponential model proposed by Mendelsohn, in which *ρ* represents a tumor growth rate constant (27). Essentially, this means that at each time interval, a fraction of tumor cells divide. The dividing fraction decreases as the tumor burden increases since substantial parts of a larger tumor mass, such as the necrotic core, are no longer able to proliferate. Since the tumor burden (T) is determined by the combination of tumor growth and tumor cell killing, the first part of Equation 1 – describing the tumor burden over time – will be:

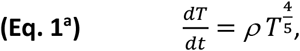

**Figure 1:**
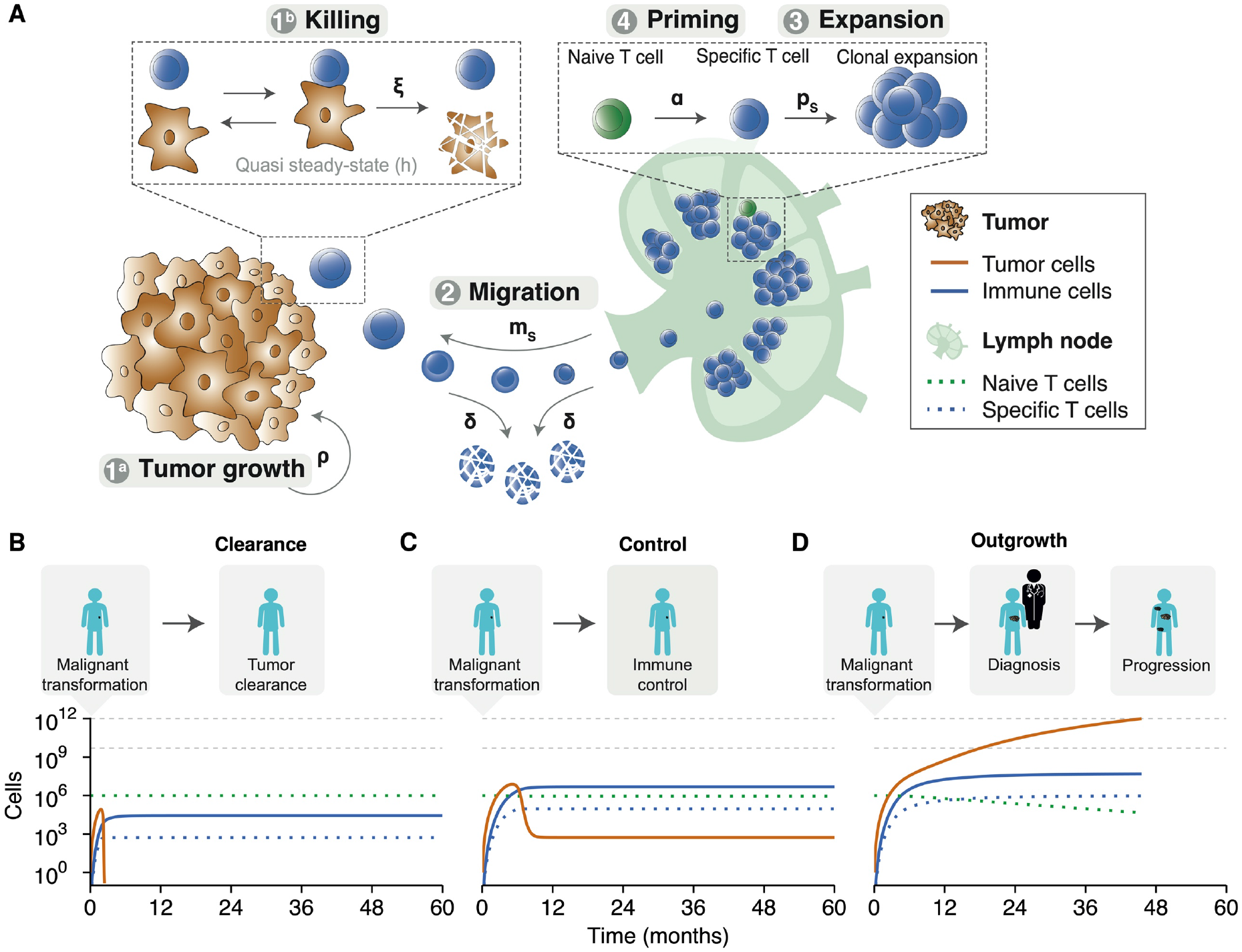
An in silico model of the tumor microenvironment generates realistic and modifiable disease courses of cancer patients. **(A)** The ODE model describes fundamental processes in the tumor microenvironment. Parameters: α = naive T cell priming rate, δ = effector T cell death rate, ξ = effector T cell killing rate, ρ = tumor growth rate, ps = effector T cell proliferation rate, and ms = effector T cell migration rate. **(B)** An effective anti-tumor immune response can eradicate tumor cells before the clinical manifestation of a tumor. **(C)** After an initial state in which the tumor outpaces the immune system, the immune system can suppress tumor growth and controls it in a subclinical state. **(D)** The natural course of disease for a clinically apparent tumor. An initial malignant transformation is followed by tumor growth until clinical diagnosis. Despite the activation of adaptive immunity, the tumor prevails. A stage of progressive disease follows, ultimately culminating in cancer-related death. The horizontal grey lines indicate (from bottom to top): the tumor burden at diagnosis and the tumor burden at death, respectively. Simulation parameters are added in Supplementary Table 1.

Where 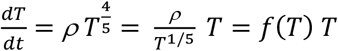, resulting in 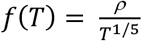 as the fraction of dividing cells per time interval, which scales inversely with the tumor burden T.

The killing rate expression is derived from the conventional Michaelis-Menten kinetics for enzyme-substrate interaction (28, 29):

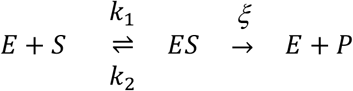

in which E, S, and P are the enzyme, substrate, and product, respectively. k_1_, k_2_, and *ξ* represent the enzyme-substrate complex formation rate, the complex dissociation rate, and the catalytic rate. Given that complex formation and dissociation occur at a rate that is at least an order of magnitude faster than tumor growth, Borghans *et al*. argued that the Michaelis-Menten kinetics could be simplified using a quasi-steady-state assumption (28). Simplification using a Padé approximation and subsequent rearrangement leads to a conventional Double Saturation (DS) model that describes effector T cell-mediated killing (28, 29):

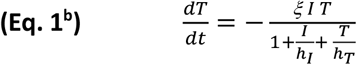

in which I is the number of immune cells in the tumor microenvironment, *ξ* is the T cell killing rate, h_I_ is the saturation constant of the effector T cells, and h_T_ is the tumor cells’ saturation constant. Here we consider T cells to follow a ‘monogamous killing’ strategy, meaning that one T cell interacts with one tumor cell at a time (28, 29).

Combining T cell-mediated tumor cell killing (Equation 1^b^) and tumor growth (Equation 1^a^), we obtain the complete differential equation that describes the tumor burden over time:

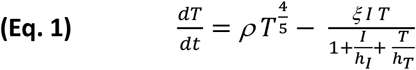

Subsequently, the immunogenicity of the tumor triggers an anti-tumor immune response. Lymph node-resident T cells (S) migrate at rate m_s_ from the lymph nodes to the tumor microenvironment. The number of intratumoral T cells over time is determined by migration and death. Therefore, by combining a migration term with a death term at rate *δ*, we obtain the following equation for the evolution of intratumoral T cells over time:

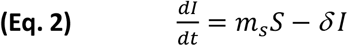

Intratumoral T cells migrate from the lymph nodes where they are produced. This process starts with converting lymph node-resident naive T cells (i.e., not activated antigen-specific; N) into antigen-specific effector T cells (S) at priming rate *α*. The priming rate *α* is scaled by the tumor size (i.e., a smaller tumor will cause less T cell priming than a larger tumor) with a scaling term 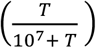, meaning that the priming rate is at half of its maximum rate and starts to saturate in tumors larger than 10^7^ cells (i.e., a sphere with a radius of 0.29 cm). Effector T cells expand clonally at proliferation rate *p*_*s*_ and migrate into the tumor microenvironment. Combining these processes, we arrive at the final two differential equations that describe the evolution of naïve and primed T cells in the lymph nodes:

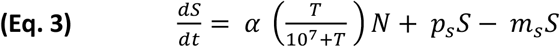

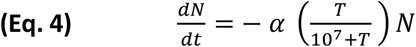

The simulations used the following initial conditions: T(0) = 1, I(0) = 0, S(0) = 0, and N(0) = 10^6^.

### Simulation parameters

The simulation parameters are listed in Table 1.

**Table 1:** Simulation parameters in the ODE model

| Symbol | Parameter (dimension) | Default value (range<sup>a</sup>) |
| --- | --- | --- |
| $\rho$ | Tumor growth rate (cells day<sup>-1</sup>) | 1 (0-7) |
| $\xi$ | Relative killing rate (cells day<sup>-1</sup>) | 0.001 (0-0.05) |
| $h$ | Michaelis constant (cells) | 571 |
| $\delta$ | Death rate of immune cells (cells day<sup>-1</sup>) | 0.019 (30) |
| $\alpha$ | Conversion rate of naive T cells into specific T cells (cells day<sup>-1</sup>) | 0.0025 |
| $p_s$ | Total production rate of effector T cells from lymph nodes (cells day<sup>-1</sup>) | 1 |
| $m_s$ | Migration rate from lymph node to tumor microenvironment (cells day<sup>-1</sup>) | 1 |

<sup>a</sup>: if not fixed

The parameters were chosen to mimic realistic *in vivo* intercellular behavior. The rationale for the choice of each parameter is explained below.

In a human adult, an estimated repertoire of approximately 10^10^ - 10^11^ naive CD8^+^ T cells is present (31, 32). Naive CD8^+^ T cells need to be primed to become activated effector T cells. The CD8^+^ T cell precursor frequency – the frequency at which any given peptide-MHC complex is recognized by naive antigen-specific CD8^+^ T cells – is on the order of 1 : 100.000 (31). Priming should be limited primarily to naive CD8^+^ T cells in one of the tumor areas draining lymph nodes. A human body contains ±600 lymph nodes. At a steady state, roughly 40% of all lymphocytes reside in lymph nodes, meaning that 40.000 naive T cells (≈ 70 naive CD8^+^ T cells per lymph node) can be primed (33, 34). We assume that priming occurs primarily in the tumor-draining lymph node station (per station harboring around 20 lymph nodes (35)). Then, 1400 T cells would be available for priming at any given time, and this pool would be refreshed approximately once per day by T cell recirculation. Considering that dendritic cells might present multiple epitopes and antigens, and that T cell priming *in vivo* might occur suboptimally, we set a priming rate of at most 2500 cells per day. The order of magnitude of these priming rates corresponds to priming rates found in chronic infectious diseases (36). Due to evasive mechanisms, anti-tumor immunity is a more dormant process than an immune response to infections (37). Therefore, we scaled the priming rate with tumor size, which translates into a maximum production of 10^6^ antigen-specific CD8^+^ T cells per day via clonal expansion. Next, we assume that all antigen-specific effector T cells migrate into the tumor microenvironment to interact with tumor cells (i.e., complex formation).

Complex formation and dissociation rates are described by the ‘Michaelis constant’, which we derived from the literature (29). The Michaelis constant describes the ratio between complex formation and dissociation.

The killing rate of effector T cells has been investigated mainly in the context of infectious disease. In their review (38), Halle *et al*. discuss discrepancies between *in vitro* and *in vivo* killing rates of effector T cells. Depending on the context, killing rates of effector T cells vary from 1 target per 5 minutes to 0-10 targets per day (38), but tumor cells are considered difficult to kill. Extensive variation in experimental *in vivo per capita* killing rates (i.e., the number of cells killed by an effector T cell per unit of time) complicates the selection of a default fixed killing parameter. Therefore, we investigated T cell dynamics over a wide range of killing rates as described using the monogamous killing regime in a double saturation model by Gadhamsetty *et al*. (29). The double saturation model ensures that the killing rate saturates with respect to the tumor cell and the effector cell densities. Consequently, our model’s maximum per capita killing rate is 2.5: one T cell can kill at most 2.5 tumor cells per day, provided there are abundant target cells available, and there is no competition with other T cells. The default tumor growth rate is one cell day^-1^, but we varied this parameter extensively in our simulations. Taken together, our default parameter values led to simulations of disease courses with realistic survival times in patients with malignancies and matched the order of magnitude of tumor growth rates as reported by others (39).

### Time-varying parameters

For the simulations shown in Figures 4 and 5, we varied the tumor growth rate *ρ* and the T cell killing rate *ξ* in a stochastic manner over time. Briefly, we set one value per month of simulated time by multiplying the baseline parameter value with a random number drawn from a normal distribution with a fixed standard deviation. The values used for the standard deviations are given in Supplementary Tables 4 and 5 (“stochasticity”). From these monthly reference values, we generated time-dependent functions using cubic B-spline interpolation. For details, see our simulation code (link given below).

### Patient simulations

We simulated tumor development in patients up to a maximum of 5 years. Note that depending on emergent tumor-immune dynamics, simulated patients may not reach the overall survival endpoint during this interval. Each time step in the simulation corresponded to one day. At baseline, one tumor cell and a pool of 10^6^ naïve tumor-specific T cells are present in a patient. Activated effector T cells are absent. We defined the time of diagnosis as the time at which the tumor exceeded 65 * 10^8^ cells and became clinically apparent. This cut-off corresponds to the assumption that a tumor with a volume of 1 cm^3^ contains 10^8^ tumor cells (40) and that several primary tumors (e.g., lung cancer, colon carcinoma, and renal cell carcinoma) are diagnosed as spherical structures with a median diameter of approximately 5 cm (41-43). The ‘lethal’ tumor burden of patients in these simulations is estimated at 10^12^ cells, corresponding to a total tumor mass of approximately 22 * 22 * 22 cm.

### Validation cohort

Model findings related to biomarker discovery programs were validated in a cohort of 58 patients with metastatic cutaneous melanoma that were treated with dendritic cell vaccination. Full details of this cohort, including baseline characteristics, were published previously (44). None of the patients received prior or subsequent immunotherapy. The serum lactate dehydrogenase levels at baseline (i.e., before therapy) were analyzed as a surrogate marker for tumor growth. The ratio of intratumoral versus peritumoral T cell densities (I/P ratio), obtained by immunohistochemical staining of the primary tumor, was selected as a surrogate marker for the T cell killing rate. Overall survival data were available for all patients.

### Model implementation

We implemented our ODE model in C++. The Boost library ‘odeint’ was used to solve the system of ordinary differential equations (45). The code is available at GitHub: https://github.com/jeroencreemers/tipping-point-cancer-immune-dynamics. Analyses and visualizations were performed in R.

## RESULTS

### Modeling tumor-immune dynamics yields realistic disease trajectories

To investigate the consequences of tumor-immune dynamics on the survival kinetics of patients, we used a computational modeling approach. We aimed to capture the interplay between tumor- and immune cells in the tumor microenvironment and simulate tumor growth in patients (see Methods). Our ODE model captured essential processes in anti-tumor immunity: priming of naive antigen-specific CD8^+^ T cells, clonal expansion of effector T cells in lymph nodes, tumor growth leading to effector T cell attraction into the tumor microenvironment, and formation of tumor-immune cell complexes to enable tumor cell killing (Fig. 1A).

We simulated tumor development from malignant transformation of a single cell, via clinical detection of a tumor, to advanced disease and possibly death. Depending on the tumor growth and the cytotoxic capacity of effector T cells, the ‘time to clinical manifestation’ and overall survival varied. Despite this variation, our simulations consistently showed three possible outcomes: 1) effector T cells inhibited tumor cell outgrowth and eradicated the tumor before clinical manifestation (Figure 1B); 2) effector T cells were initially unable to inhibit tumor cell outgrowth but caught up and suppressed tumor growth to a balanced subclinical state (Figure 1C); or 3) exponential tumor growth outpaced the immune system’s control and gave rise to a clinically detectable tumor (Figure 1D). These three scenarios only led to two clinically different outcomes in patients: either a tumor became clinically evident, or the immune system could suppress or eradicate a tumor at an early stage (i.e., before the tumor could reach a clinically detectable size). A balanced equilibrium state, in which the immune system keeps a clinically evident tumor under persistent control, does not exist in this deterministic version of our model.

### Patient survival depends on a tipping point in tumor-immune dynamics

To better characterize these dichotomous survival kinetics, we examined how tumor-immune dynamics influenced patient survival by varying the tumor growth rate and the T cell killing rate over a broad range of possible values.

First, we focused solely on the tumor-component by varying the tumor growth rate. An increase in tumor growth did not gradually shorten overall survival in patients (Figure 2A). On the contrary, a critical threshold was present. Once the threshold was exceeded, the kinetics ‘flipped’ from a state of immune control (Figure 2A, inset 1) to a state in which the tumor could evade immune control (Figure 2A, inset 2).

**Figure 2:**
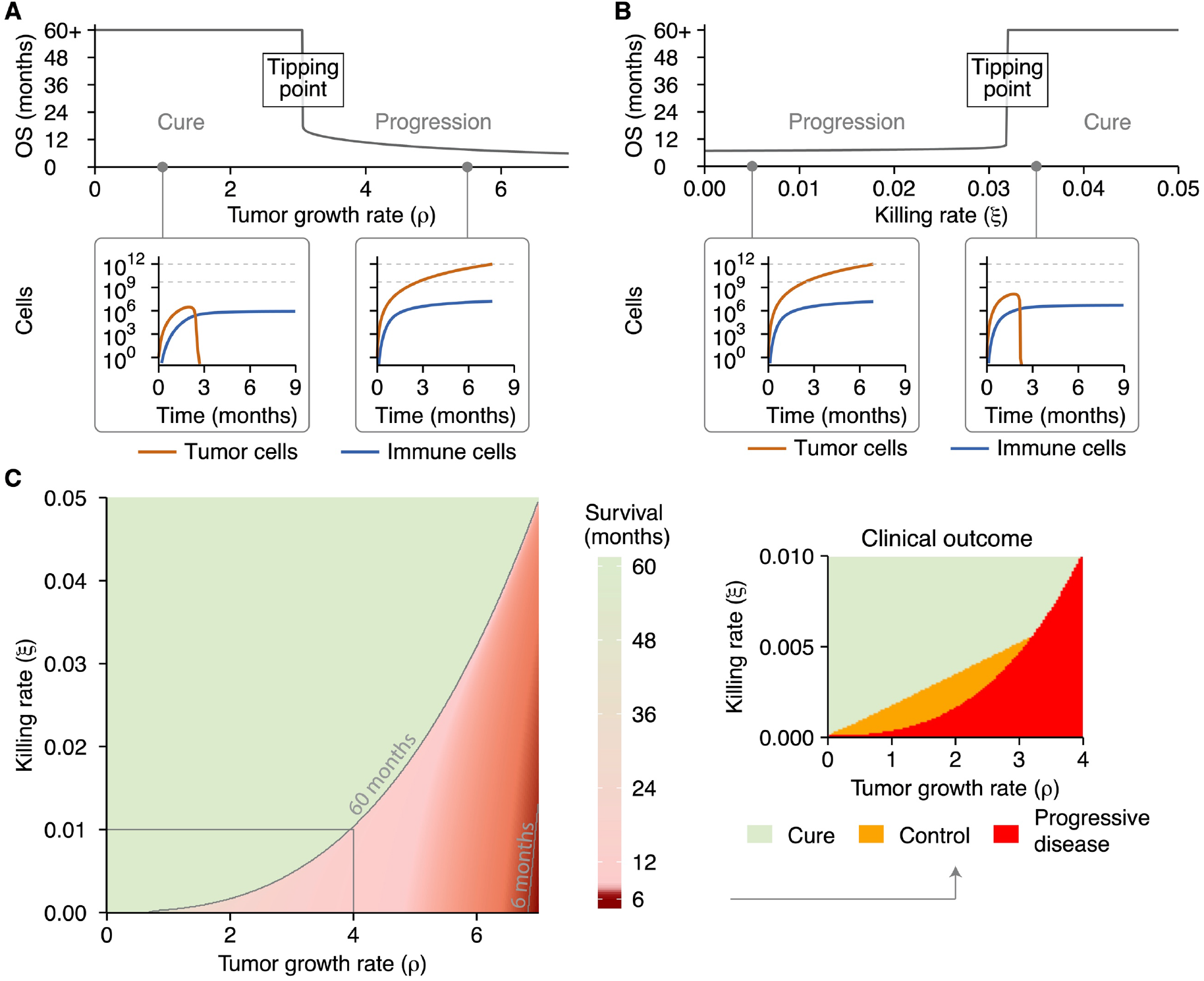
A tipping point in the tumor-immune interaction determines a patient’s outcome. **(A)** A gradual increase in tumor growth reveals a tipping point, where long-term survival (immune control; inset 1) abruptly changes to short-term survival (immune evasion; inset 2). **(B)** A similar analysis reveals a tipping point along the immune axis, again differentiating short-term survival (immune evasion; inset 1) from long-term control (immune control; inset 2). **(C)** The tipping point is present across the entire range of parameters examined. Cure and progressive disease are the dominant states, whereas subclinical tumor control only occurs within a limited parameter range (inset). Simulation parameters are shown in Supplementary Table 2.

Second, we investigated the influence of the T cell killing rate on overall survival. As for the death rate, a gradual increase in the cytotoxic capacity of effector T cells did not induce a gradual change in survival times. Instead, a sharp state transition that differentiated short from long survival was observed again (Figure 2B). This coincided with the phenotypes ‘immune evasion’ (Figure 2B, inset 1) and ‘immune control’ (Figure 2B, inset 2).

To visualize this sudden state transition or ‘tipping point’ in tumor-immune dynamics as a function of both tumor proliferation and cytotoxic killing at the same time, we visualized the joint influence of the tumor growth rate and T cell killing rate on survival in a heatmap (Figure 2C). This ‘phase diagram’ shows that the tipping point is not only present for specific parameter values but is a fundamental property in our model. By contrast, the state of subclinical tumor control was not universally present around the tipping point (Figure 2C, inset) but manifested itself only in a narrow range of parameters. Within both the ‘Cure’ and ‘Control’ domain (Figure 2, inset), the immune system prevented tumors from reaching a detectable size, precluding the clinical classification as ‘patient’. The difference between individuals in the ‘Cure’ and ‘Control’ domains was that all tumor cells were eradicated in the former, while in the latter, the immune system kept the tumor in an undetectable subclinical state (i.e., a tumor size of around 10^3^ tumor cells; Figure 1B/C).

Next, we expanded these analyses to characterize the tipping point in different tumor types. A fundamental distinction between tumors is the rate at which they induce T cell priming, for instance, through tumor-specific immunogenicity or by specific characteristics of the immunosuppressive microenvironment. To this end, we simulated four tumor types: a tumor without T cell priming and three tumors in which T cell priming was varied from low to high. Without T cell priming, survival was only determined by the tumor growth rate – logically, no tipping point exists in the absence of T cells (Supplementary Figure 1A). With T cell priming, tipping points became apparent. The location of the tipping point was affected by the priming rate. A higher priming rate facilitated improved tumor eradication through an increased influx of cytotoxic T cells into the tumor microenvironment (Supplementary Figure 1B-D).

In general, the presence of a tipping point indicates that small perturbations in either tumor growth rate or T cell killing rate in the vicinity of a tipping point may result in substantial overall survival differences in patients. In contrast, much larger perturbations far away from the tipping point would have far less effect.

### Immune checkpoint inhibitors induce a survival benefit by shifting patients over a tipping point

So far, we have described tumor-immune interactions during the natural course of malignant disease. In a clinical setting, however, therapeutic interventions are available to steer disease courses.

Dependent on the treatment of choice, a specific effect is exerted on the tumor microenvironment. Treatment effects vary from constraining the proliferative capacity of tumor cells (e.g., chemotherapy or targeted therapy) to increasing the T cell pool (e.g., CAR T cells) or expanding the proliferative capacity of T cells (e.g., cancer vaccines; Figure 3A). Given the unparalleled responses of advanced malignancies to immunotherapy, we focused on the consequences of a tipping point for responses to immune checkpoint inhibitors (ICI), but these findings could be extended to other therapies as well. In this study, we limited the treatment effect of ICI to their primary mode of action: the augmentation of the T cell killing rate (Figure 3A).

**Figure 3:**
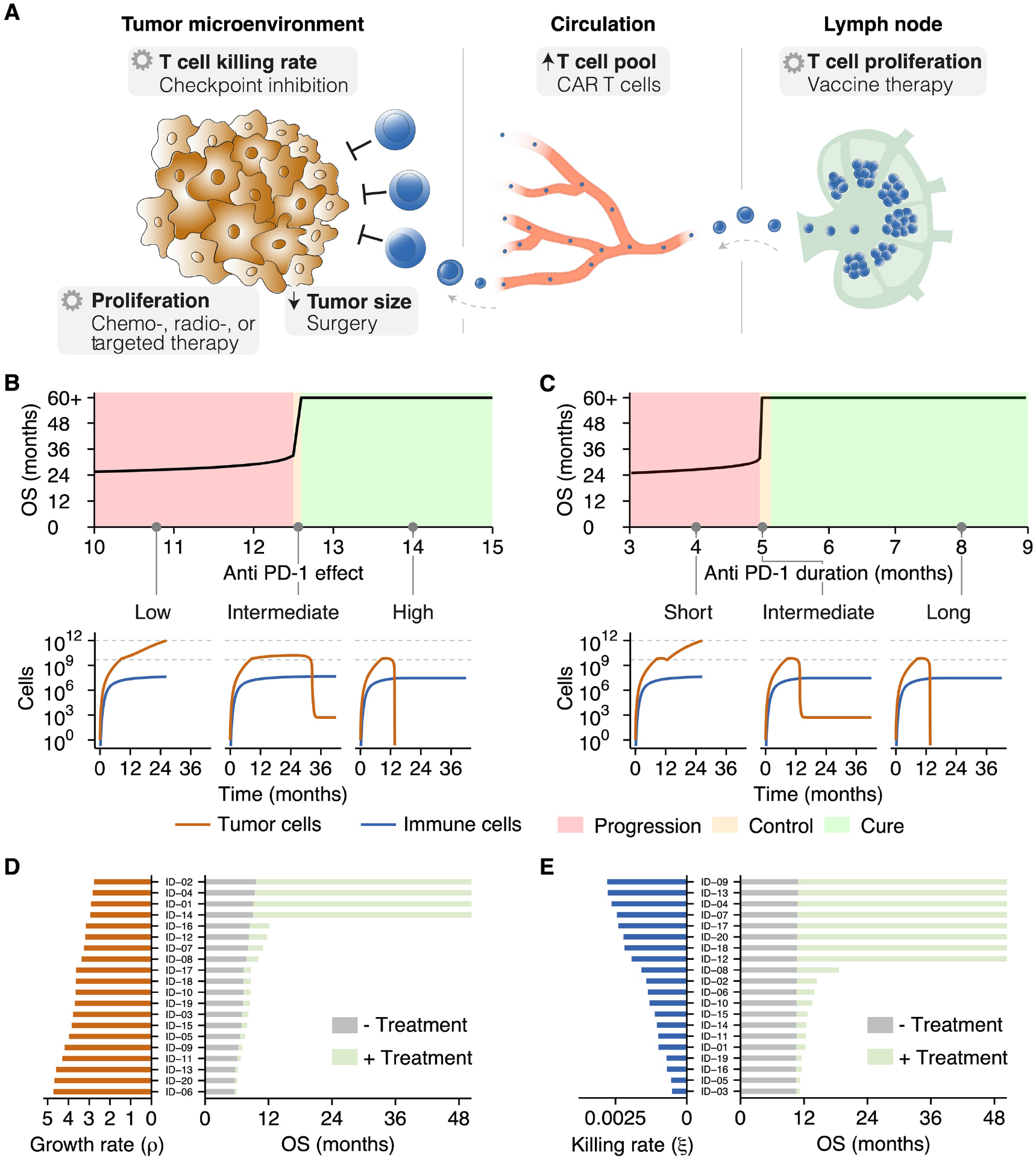
Tipping points induce dichotomous clinical outcomes in heterogeneous patient populations. **(A)** Treatments target processes or cell populations in the tumor microenvironment. **(B-C)** Two criteria need to be met to induce long-term survival: **(B)** ICI need to augment T cell killing sufficiently, and **(C)** the treatment effect needs to be retained for a prolonged time. An inadequate treatment effect or limited treatment duration led at maximum to a temporary survival benefit. (D-E) In patient populations with variation in only **(D)** the tumor (i.e., growth rate), or **(E)** the immune system (i.e., T cell killing rate), the distance to a tipping point determines the clinical benefit. Without treatment, survival was limited (grey bars). In contrast, ICI induced long-term survival solely in patients close to a tipping point (green bars). See also Supplementary Table 3.

**Figure 4:**
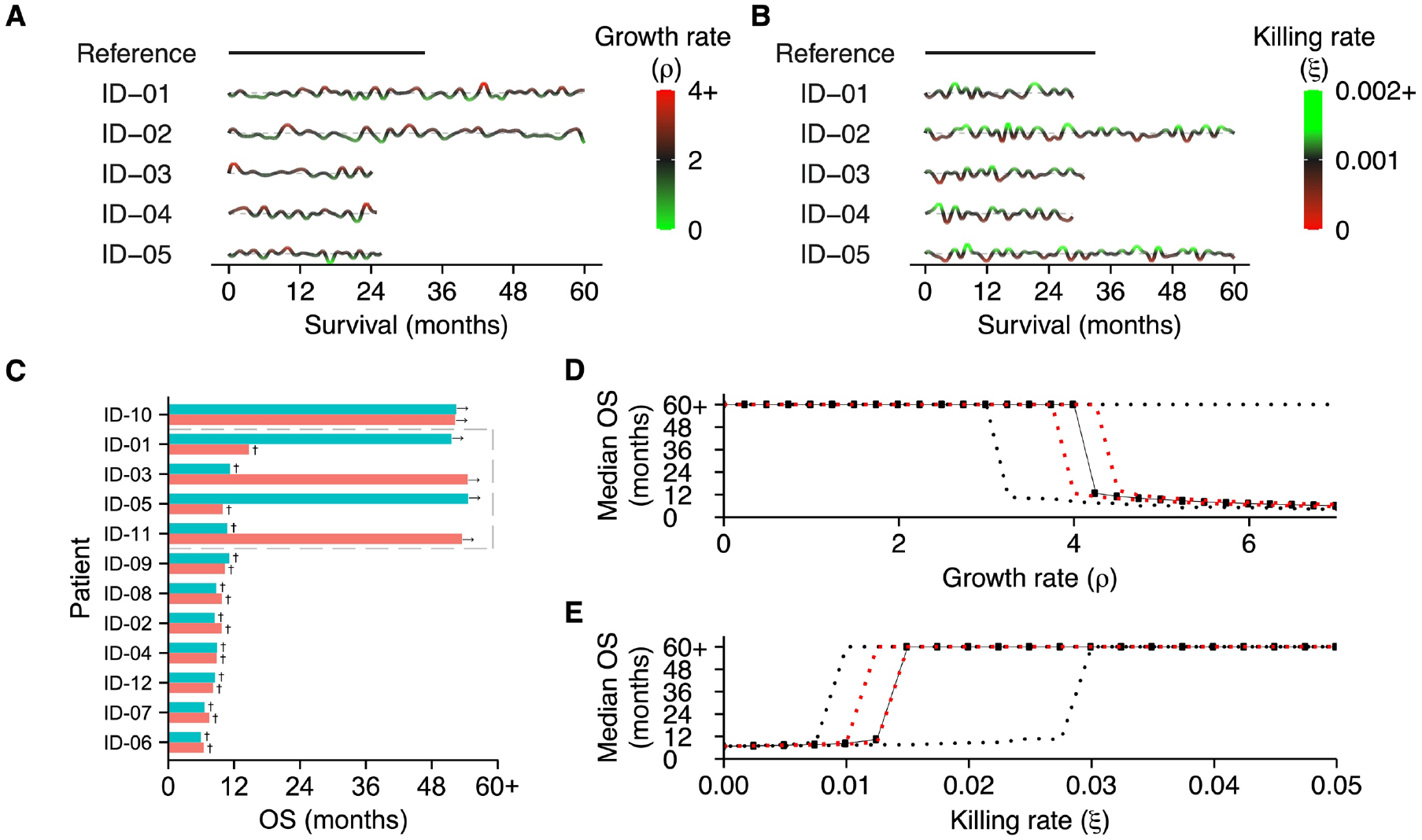
Survival outcomes are strongly affected by evolving patient dynamics. **(A-B)** Examples of dynamic disease courses in patients with identical tumors and immune systems at baseline, respectively. **(A)** Evolving tumors (i.e., random variation in tumor growth rate over time) and **(B)** continuous variation in the potency of the immune system (i.e., killing rate) lead to divergent survival outcomes. The grey dotted lines indicate the baseline values for the growth rate and killing rate, respectively. **(C)** Dynamic trajectories in a heterogeneous patient population can move patients towards or away from a tipping point. The grey box indicates patients in which dynamic trajectories strongly alter survival outcomes. See also Supplementary Figure 2. In dynamic trajectories, **(D)** baseline tumor growth and **(E)** baseline T cell killing rates cannot accurately predict overall survival. Note: all patients in these examples are treated with ICI. The red and black dotted lines indicate the 25% and 75% quantiles, respectively. See also Supplementary Table 4.

**Figure 5:**
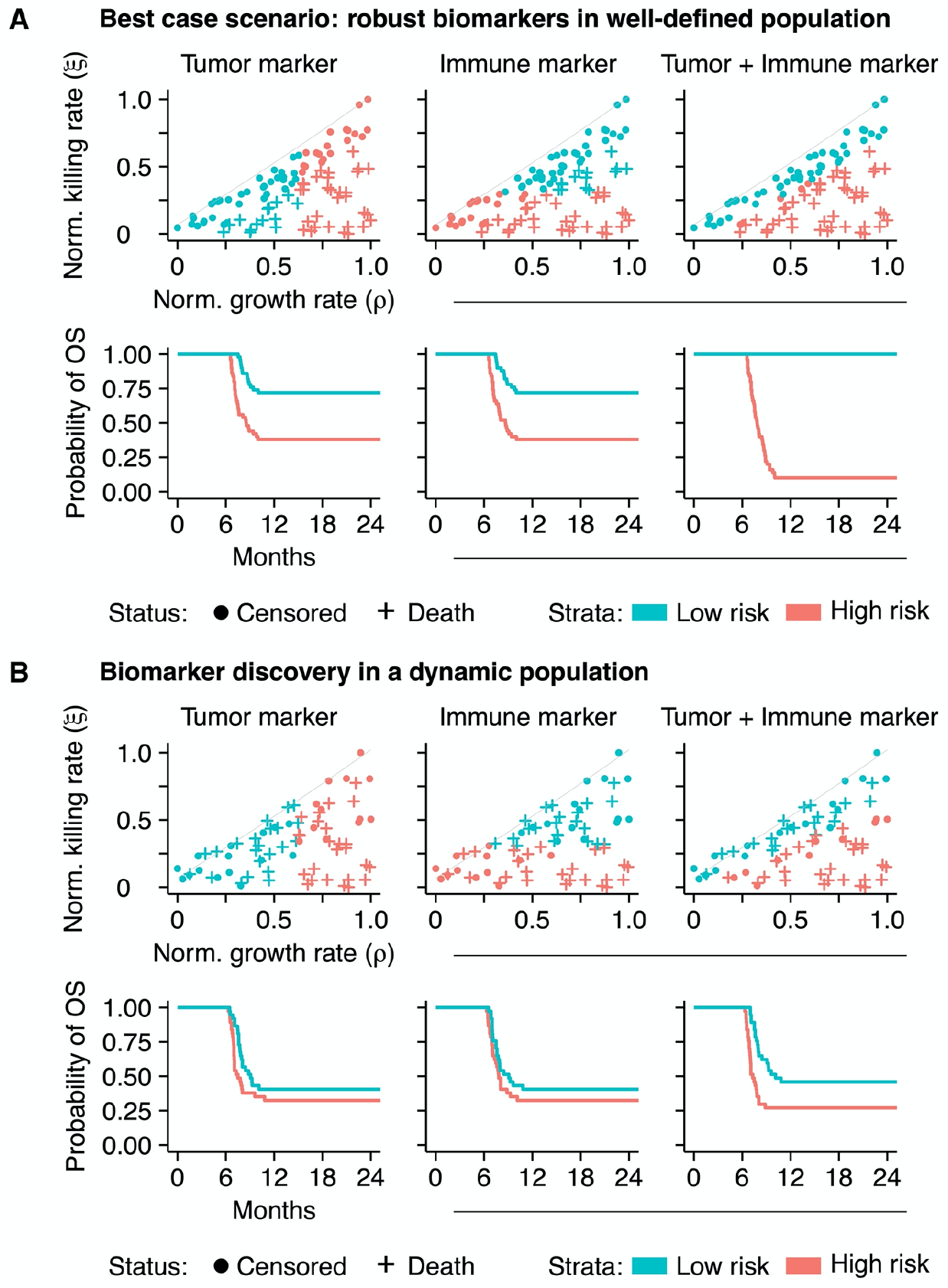
Non-linear tumor-immune dynamics complicate biomarker discovery. **(A)** An in silico biomarker discovery study in a ‘fixed’ patient cohort: while a single biomarker – either a tumor or an immune marker – can predict survival to some extent (the first and second columns), information from both markers in a biomarker panel enhances the predictive capacity greatly (third column). **(B)** Dynamic disease trajectories challenge survival prediction with ‘baseline’ biomarkers. In dynamic disease courses, the predictive value of single ‘baseline’ biomarkers is limited (the first and second columns; compare to Figure 5A). A biomarker panel improves survival predictions in this cohort (the third column) but is still defied by evolving dynamics. See also Supplementary Table 5.

In the presence of a tipping point, ICI could induce a long-term survival benefit under two conditions: 1) the effect of treatment needs to be potent enough to shift a patient over a tipping point (Figure 3B), and 2) the treatment effect needs to be sustained long enough for a patient to benefit from the treatment (Figure 3C). The treatment effect was defined as the multiplication factor of the T cell killing rate. When both criteria were satisfied, ICI were able to induce a long-term survival benefit. However, if the treatment effect (anti-PD1 effect < 12.6) or duration (less than ±5 months) proved inadequate, any survival benefit was only temporary, and inevitable tumor progression would ultimately limit overall survival (Figure 3B/C; insets). These survival kinetics depend not solely on therapeutic features of ICI but rather on the interplay between patient and ICI characteristics. To illustrate this, we simulated twenty patients with identical immune systems (i.e., identical T cell killing rates). In the absence of ICI therapy, variation in the tumor growth rate – that is, variation in the distance to a tipping point – led to a limited variation in survival (Figure 3D; grey bars). When these same patients were treated with ICI, a survival benefit is induced in all patients. However, the extent of this benefit differs and depends on the distance to a tipping point. Following clinical observations, long-term survival is only induced in the subset of patients close to a tipping point (Figure 3D; green bars). Similar findings were obtained in a population of patients with identical tumors but different immune systems. Without treatment, hardly any survival variation is present (Figure 3E; grey bars). Again, treatment with ICI induced dichotomous clinical outcomes: a small survival benefit in most patients, with long-term survival in a subset (Figure 3E; green bars). Hence, the mere presence of a tipping point yields heterogeneity in treatment outcomes.

### Tipping points determine patient outcomes in dynamic patient trajectories

Thus far, our simulations considered tipping points generated in patients with fixed characteristics. However, disease courses in patients are certainly not fixed and are, to a certain extent, subject to (possibly random) variation. We hypothesized that interpatient variability in clinical outcomes could be partially attributable to this dynamic behavior of cancers and the interaction with the immune system. Such variation might reflect biological processes (e.g., accumulating mutations, the expression of checkpoint molecules, and the availability of nutrients) that alter anti-tumor immunity and promote or hamper tumor development. We reasoned that the subsequent dynamics could drive patients towards and ultimately over a tipping point – or move patients away from it, which would limit the survival benefit of these treatments. To verify this hypothesis, we simulated the effect of dynamically evolving tumors (Figure 4A) or immune systems (Figure 4B) in identical patients compared to a static reference patient. Specifically, we varied the tumor growth rate and the T cell killing rate randomly over time (parameter values are included in Supplementary Table 4). Upon reaching a diagnosable tumor volume, all patients in these examples were treated with ICI. As expected, stochastic dynamics prompted survival differences and induced a survival benefit in a subset of patients. In a heterogeneous patient population, this led to an interesting finding: the initial distance to a tipping point, along with the dynamics itself, determined the clinical outcome of patients treated with ICI (Figure 4C, Supplementary Figure 2). At population level, this led to a distinction between three subsets of patients: (1) patients far away from a tipping point with an unmodifiable bad prognosis (non-responders), (2) patients close to a tipping point with a favorable prognosis (responders), and, most importantly, (3) patients in between these groups (potential responders). In the last subset, tumor dynamics ultimately determined the treatment response, and thereby the clinical outcome (Figure 4C; grey box). A clinically important ramification of dynamic trajectories is that even if the subset to which a patient belongs is known at baseline, dynamics could alter the distance to a tipping point and, thereby, the prognosis of a patient. Therefore, it might be impossible to predict the prognosis solely based on characteristics measured upon diagnosis. Dynamic trajectories can significantly diversify patient outcomes, meaning that continuous variation in the tumor growth rate (Figure 4D) or T cell killing rate (Figure 4E) leads to an entire spectrum of patient outcomes.

### Implications of tipping points for biomarker discovery studies

Biomarker discovery studies aim to improve the prediction of patient survival upon treatment. We observed that tipping points are crucial in shaping survival kinetics. Therefore, accurate survival predictions would require the consideration of tipping points. Ideally, a prognostic biomarker (or biomarker panel) would consistently distinguish long-term survivors from their counterparts. Since the non-linear survival dynamics following a tipping point weaken the correlation between a single biomarker and survival, the question is: how can we screen for biomarkers in a more efficient manner that takes this tipping point into account?

At first, we approached this question with an *in silico* biomarker discovery study. We measured the value of two potential biomarkers at baseline in simulated patients (n=100) that were subsequently treated with ICI (cohort characteristics are specified in Supplementary Table 5). We simplified the cohort by fixing the tumor and immune characteristics of these patients over time and assumed to have access to an entirely accurate biomarker (i.e., no measurement error; Figure 5A). Within this cohort, we predicted the prognosis of patients based on either the tumor or immune marker (the first and second columns of Figure 5A, respectively). As is common in practice (though from a statistical point of view far from ideal), we dichotomized the biomarker using its median as a cut-off. Although survival differentiation based on these biomarkers alone was partially possible, it remained far from optimal. However, when we constructed a biomarker panel including both biomarkers, it highly accurately discriminated short-term from long-term survivors (third column of Figure 5A). Note that despite variability in time from diagnosis, the initial plateau in the survival curves was caused by the fact that all tumors were diagnosed with identical sizes and immediately treated.

In clinical practice, the assumption of a ‘fixed’ patient trajectory does not hold. Therefore, we simulated this cohort again with dynamic trajectories. Due to the dynamics, a subgroup of patients did not develop clinical tumors and was excluded from the analysis. The prediction of a patient’s prognosis with a single biomarker, either from the tumor or the immune system, in a dynamic cohort became increasingly challenging (the first and second columns of Figure 5B). The combination of both markers in a biomarker panel increased the predictive capacity slightly, enabling the prediction of prognosis to some extent. However, in line with the notion of personalized medicine, the accurate and individualized prediction of prognosis based on baseline characteristics was not feasible in a significant subgroup of patients due to dynamic tumor-immune interactions (third column of Figure 5B).

These *in silico* experiments suggest that biomarker discovery efforts benefit from considering tumor and immune markers in concert rather than alone. To test this hypothesis, we retrospectively analyzed clinical data derived from previous trials in patients with metastatic melanoma (n = 58; see baseline characteristics in Supplementary Table 6) (44). We assessed whether a combination of two biomarkers would provide more information on a patient’s survival than either marker alone. Baseline lactate dehydrogenase (LDH) was selected as a surrogate marker for tumor growth, and the ratio between immunohistochemically-determined intratumoral vs. peritumoral immune cells (I/P ratio) on the primary tumor was selected as an immune marker. We then used two different methods to measure the amount of information these markers provide on patient survival. First, we applied linear discriminant analysis to determine marker cut-off values that distinguish “short survivors” (<9 months) from “long survivors” (>9 months, corresponding to the median survival in the cohort). A cut-off based on the tumor marker LDH alone correctly classifies 71% of patients (Figure 6A), which increased to 78% when using the I/P ratio as an immune marker instead. A combination of both markers achieves 86% accuracy, with the discrimination line following a roughly diagonal slope akin to the tipping point in our “in silico” cohort (Figure 2).

**Figure 6:**
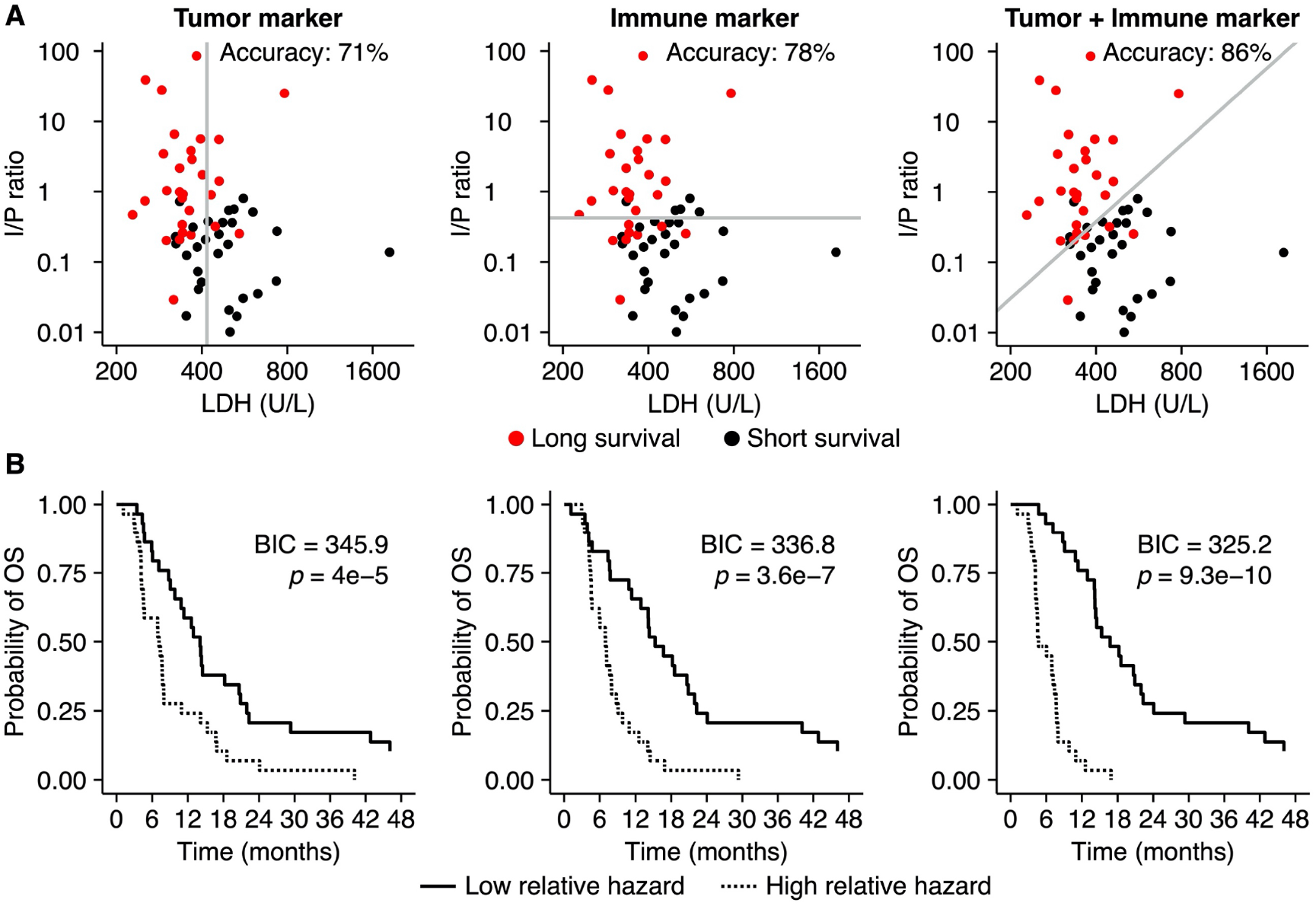
**A composite biomarker consisting of a tumor and an immune component outperforms single markers in a retrospective analysis of metastatic melanoma patients.** **(A)** A linear classifier based on LDH level at baseline, a surrogate marker for tumor growth, classified 71% of all patients correctly as short survivors (<9 months) or long survivors (>9 months). With an accuracy of 78%, the I/P ratio – an immune marker – performs better in this cohort. A linear combination of both markers leads to an even better classification (86% accuracy) than either one alone. **(B)** A Cox proportional hazard model based on both markers fits the data better than models based on either marker as measured by the Bayesian Information Criterion (BIC) (the lower, the better, and differences above 10 are considered strongly favoring one model over another).

Second, we compared Cox proportional hazard models based on LDH alone and I/P ratio alone to a model including both markers. Both LDH (likelihood ratio test: p=4×10^−7^) and I/P ratio (p=3.6×10^−7^) explained survival better than chance on their own, but a bivariable model (p=9.3×10^−10^; Supplementary Table 7) provided the best fit to the data as measured by a Bayesian Information Criterion (BIC), which was lower by 11.6 compared to the LDH-only model and by 20.7 compared to the I/P ratio-only model. The Kaplan-Meier plots shown in Figure 6B illustrate the performance of each model by comparing the patients with the highest 50% estimated relative hazard to the lowest 50%. These results support our *in silico*-generated hypothesis that a combination of tumor and immune markers form a better basis for patient stratification than either marker on its own.

Two important findings are derived from these observations: First, due to the non-linear tumor-immune dynamics with respect to survival, it can be complicated for a single biomarker to predict a patients’ prognosis accurately. Since survival kinetics emerge from the interplay between a cancer and the immune system, biomarkers from both systems need to be incorporated simultaneously into a biomarker panel to improve the predictive value. Second, biomarker measurements at baseline are merely a situational snapshot of the disease conditions *at a specific point in time*. Depending on the magnitude of the dynamics, it might become challenging or even impossible to predict the prognosis of patients from these biomarkers correctly.

## DISCUSSION

This study investigated how tumor-immune dynamics relate to ICI-induced treatment responses and survival kinetics of patients. We predict that a tipping point is present in the tumor-immune interaction. This finding implies that underneath the intricate interplay between a developing malignancy and the immune system, two contrasting disease states determine disease outcome: a state where the immune system controls tumor outgrowth and a state in which a tumor escapes immune defense. A stable “steady state” in which tumor growth and the immune response perfectly balance each other for extended periods seems only plausible in a subclinical setting. We show that treatment with ICI can induce a survival benefit by shifting a patient over a tipping point, thereby tipping the balance in tumor-immune dynamics in favor of survival. In line with clinical observations of interpatient variability in disease courses, we found that dynamics in patient trajectories pose major challenges for treatment response prediction. Moreover, we showed how the tipping point in dynamic patient trajectories defies simple strategies for outcome prediction in biomarker discovery studies. In particular, when facing highly dynamic disease courses, adaptive treatment strategies based on continuous monitoring might be more promising than simple patient stratification at baseline.

Tipping points are well-known in complex systems such as financial markets and ecosystems but are also present in medicine (46, 47). State transitions might progress gradually or abruptly. If a system balances around a critical threshold, small perturbations might induce an abrupt transition to a contrasting state. In oncology, phenomena like partial or complete radiologic responses during treatment or (hyper)progression after discontinuation of treatment suggest the presence of state transitions (48, 49). Based on these observations, a tipping point in cancer immunotherapy had been speculated upon (50). Experimentally, tipping points are most clearly represented by early preclinical work in the PD-1/PD-L1 axis. Consistent with our findings, dichotomous treatment responses arise in syngeneic DBA/2 mice inoculated with P815/PD-L1 cells (51). While genetically identical with similar tumor characteristics, anti-PD-L1 antibodies prolong survival in only a subset of the mice, likely due to stochastic differences in immune responses and TCR repertoire. Additional *in vivo* data supporting the theory of tipping points in oncology is derived from studies on dynamic network biomarkers, showing its relevance during the onset of metastasis in hepatocellular carcinoma (52) and the development of treatment resistance in breast cancer (53). This study provides a potential mechanistic explanation for this phenomenon in immuno-oncology and shows its implications on the induction of long-term survival in clinical practice and biomarker discovery. From a biomechanistic perspective, such state transitions in cancer immunotherapy arise due to fundamental differences in proliferation kinetics between tumors and the immune system. While tumor cell proliferation is virtually unrestricted, immune cell proliferation is much more limited and tightly controlled. Our finding that tipping points affect not only natural disease courses but also treatment responses underlines the importance of these kinetics.

Tipping points within tumor-immune dynamics have important implications for biomarker discovery. Biomarkers are developed to predict prognosis and steer clinical decision-making. Disease outcomes in cancer patients are essentially determined by the interplay between two complex systems: the tumor and the immune system. Our model predicts that factors from both systems should be considered to improve the predictive power of biomarkers. However, in contrast with this seemingly straightforward prediction, current research mainly focuses on factors derived from one of the two complex systems. Expression of programmed death ligand-1 (PD-L1) on tumor tissue illustrates this: while 45% of patients with PD-L1 positive tumors show objective responses to anti-PD(L)1 immunotherapy, 15% of patients with PD-L1 negative tumors also show objective responses (54). Other explanations for this difference include heterogeneous intratumoral and inter-metastases expression patterns, positivity-threshold selection, and differences in immunohistochemical staining protocols. In that respect, tumor mutational burden (TMB) might prove to be a highly relevant biomarker. The mutation rate is a tumor-intrinsic factor associated with the phenotypical aggressiveness of tumors (55). Simultaneously, a high mutational burden might induce a plethora of neoantigens, linking this tumor-intrinsic factor directly to adaptive immunity. Clinical observations of a stronger association between TMB and response rates to anti-PDL1 immunotherapy compared to PD-L1 expression in patients with urothelial carcinoma support this hypothesis (56). Our research thus reinforces common calls to integrate multiple biomarkers for immunotherapy prediction outcomes (57, 58); at least, a combination of both immunological and tumor-related parameters should be the basis of *any* biomarker discovery effort. The strongly non-linear dynamics resulting from the tipping point mean that a one-dimensional approach will likely be insufficient.

Our approach has to be interpreted in light of some limitations. Although the ‘coarse-grained’ nature of ODE models allows focusing on the major common underlying mechanisms in many cancers, it is also a potential pitfall. For example, metabolic processes such as hypoxia, immune-suppressive characteristics of the tumor microenvironment such as the presence of FoxP3^+^ regulatory T cells or expression of transforming growth factor β, the presence of other relevant effector cells such as natural killer cells, and the availability of nutrients are only implicitly represented by our model in a single killing efficacy parameter. This simplification also holds for treatments. In this study, ICI was limited to its main mode of action: the augmentation of the T cell killing rate. While the ‘true’ mechanistic effects might be more widespread, sufficient data to correctly parameterize more complex models remains scarce. Furthermore, it should be emphasized that an ODE model contains limited spatial information; while we distinguish between lymphatic tissue and the tumor microenvironment, all cells within the microenvironment are identical, and all processes affect cells in the same manner. Although we do not expect that explicit incorporation of these processes or translation of the model into a spatial variant alters our central finding of a tipping point, it could nevertheless be of interest to verify these hypotheses in future research using more complex, spatial agent-based models.

In conclusion, we used computational modeling to show that the clinical outcome of cancer patients is determined by tipping points in tumor-immune dynamics. A tipping point influences not only treatment response but also the prognosis of patients and has major implications for future biomarker research.

## Data Availability

The code of the ODE model is available at GitHub.

https://github.com/jeroencreemers/tipping-point-cancer-immune-dynamics.

## LIST OF ABBREVIATIONS

BIC: Bayesian Information Criterion
ICI: Immune Checkpoint Inhibition
LDH: Lactate dehydrogenase
ODE: Ordinary Differential Equation
PD-(L)1: Programmed Death-(Ligand) 1
TMB: Tumor Mutational Burden

## DECLARATIONS

### Ethics approval and consent to participate

Not applicable.

### Consent for publication

Not applicable.

### Availability of data and material

The code of the ODE model is available at GitHub: https://github.com/jeroencreemers/tipping-point-cancer-immune-dynamics.

## Authors’ contribution

JHAC and JT conceived this study. JHAC performed the experiments and wrote the manuscript under the supervision of JT. All authors provided feedback on the manuscript and reviewed the manuscript prior to submission.

## Acknowledgments

Not applicable.

## REFERENCES

1. Larkin J, Chiarion-Sileni V, Gonzalez R, et al. Five-Year Survival with Combined Nivolumab and Ipilimumab in Advanced Melanoma. N Engl J Med. 2019;381(16):1535–46.

2. Hellmann MD, Paz-Ares L, Bernabe Caro R, et al. Nivolumab plus Ipilimumab in Advanced Non-Small-Cell Lung Cancer. N Engl J Med. 2019;381(21):2020–31.

3. Motzer RJ, Escudier B, McDermott DF, et al. Survival outcomes and independent response assessment with nivolumab plus ipilimumab versus sunitinib in patients with advanced renal cell carcinoma: 42-month follow-up of a randomized phase 3 clinical trial. J Immunother Cancer. 2020;8(2).

4. Aly A, Mullins CD, Hussain A. Understanding heterogeneity of treatment effect in prostate cancer. Curr Opin Oncol. 2015;27(3):209–16.

5. Kent DM, Alsheikh-Ali A, Hayward RA. Competing risk and heterogeneity of treatment effect in clinical trials. Trials. 2008;9:30.

6. Bedard PL, Hansen AR, Ratain MJ, et al. Tumour heterogeneity in the clinic. Nature. 2013;501(7467):355–64.

7. Dagogo-Jack I, Shaw AT. Tumour heterogeneity and resistance to cancer therapies. Nat Rev Clin Oncol. 2018;15(2):81–94.

8. Sharma P, Hu-Lieskovan S, Wargo JA, et al. Primary, Adaptive, and Acquired Resistance to Cancer Immunotherapy. Cell. 2017;168(4):707–23.

9. Roskoski R, Jr. A historical overview of protein kinases and their targeted small molecule inhibitors. Pharmacol Res. 2015;100:1–23.

10. Kern SE. Why your new cancer biomarker may never work: recurrent patterns and remarkable diversity in biomarker failures. Cancer Res. 2012;72(23):6097–101.

11. Murphy H, Jaafari H, Dobrovolny HM. Differences in predictions of ODE models of tumor growth: a cautionary example. BMC Cancer. 2016;16:163.

12. Edelman EJ, Guinney J, Chi JT, et al. Modeling cancer progression via pathway dependencies. PLoS Comput Biol. 2008;4(2):e28.

13. Baratchart E, Benzekry S, Bikfalvi A, et al. Computational Modelling of Metastasis Development in Renal Cell Carcinoma. PLoS Comput Biol. 2015;11(11):e1004626.

14. De Mattos-Arruda L, Vazquez M, Finotello F, et al. Neoantigen prediction and computational perspectives towards clinical benefit: recommendations from the ESMO Precision Medicine Working Group. Ann Oncol. 2020;31(8):978–90.

15. Pujana MA, Han JD, Starita LM, et al. Network modeling links breast cancer susceptibility and centrosome dysfunction. Nat Genet. 2007;39(11):1338–49.

16. Friberg LE, Henningsson A, Maas H, et al. Model of chemotherapy-induced myelosuppression with parameter consistency across drugs. J Clin Oncol. 2002;20(24):4713–21.

17. Lee JJ, Huang J, England CG, et al. Predictive modeling of in vivo response to gemcitabine in pancreatic cancer. PLoS Comput Biol. 2013;9(9):e1003231.

18. Wang HW, Milberg O, Bartelink IH, et al. In silico simulation of a clinical trial with anti-CTLA-4 and anti-PD-L1 immunotherapies in metastatic breast cancer using a systems pharmacology model. Roy Soc Open Sci. 2019;6(5).

19. Valentinuzzi D, Simoncic U, Ursic K, et al. Predicting tumour response to anti-PD-1 immunotherapy with computational modelling. Phys Med Biol. 2019;64(2):025017.

20. Sun X, Bao J, Shao Y. Mathematical Modeling of Therapy-induced Cancer Drug Resistance: Connecting Cancer Mechanisms to Population Survival Rates. Sci Rep. 2016;6:22498.

21. Zhang J, Cunningham JJ, Brown JS, et al. Integrating evolutionary dynamics into treatment of metastatic castrate-resistant prostate cancer. Nat Commun. 2017;8(1):1816.

22. Cunningham JJ, Brown JS, Gatenby RA, et al. Optimal control to develop therapeutic strategies for metastatic castrate resistant prostate cancer. J Theor Biol. 2018;459:67–78.

23. Ribba B, Boetsch C, Nayak T, et al. Prediction of the Optimal Dosing Regimen Using a Mathematical Model of Tumor Uptake for Immunocytokine-Based Cancer Immunotherapy. Clin Cancer Res. 2018;24(14):3325–33.

24. Knight-Schrijver VR, Chelliah V, Cucurull-Sanchez L, et al. The promises of quantitative systems pharmacology modelling for drug development. Comput Struct Biotechnol J. 2016;14:363–70.

25. Fassoni AC, Baldow C, Roeder I, et al. Reduced tyrosine kinase inhibitor dose is predicted to be as effective as standard dose in chronic myeloid leukemia: a simulation study based on phase III trial data. Haematologica. 2018;103(11):1825–34.

26. Clark RE, Polydoros F, Apperley JF, et al. De-escalation of tyrosine kinase inhibitor therapy before complete treatment discontinuation in patients with chronic myeloid leukaemia (DESTINY): a non-randomised, phase 2 trial. Lancet Haematol. 2019;6(7):e375–e83.

27. Mendelsohn ML. Cell proliferation and tumor growth. Oxford: Blackwell Scientific Publications. 1963.

28. Borghans JA, de Boer RJ, Segel LA. Extending the quasi-steady state approximation by changing variables. Bull Math Biol. 1996;58(1):43–63.

29. Gadhamsetty S, Maree AF, Beltman JB, et al. A general functional response of cytotoxic T lymphocyte-mediated killing of target cells. Biophys J. 2014;1068):1780–91.

30. McDonagh M, Bell EB. The survival and turnover of mature and immature CD8 T cells. Immunology. 1995;844):514–20.

31. Jenkins MK, Chu HH, McLachlan JB, et al. On the composition of the preimmune repertoire of T cells specific for Peptide-major histocompatibility complex ligands. Annu Rev Immunol. 201028:275–94.

32. Coulie PG, Karanikas V, Lurquin C, et al. Cytolytic T-cell responses of cancer patients vaccinated with a MAGE antigen. Immunol Rev. 2002188:33–42.

33. Westermann J, Pabst R. Distribution of lymphocyte subsets and natural killer cells in the human body. Clin Investig. 1992;707):539–44.

34. Ferrer R. Lymphadenopathy: differential diagnosis and evaluation. Am Fam Physician. 1998;586):1313–20.

35. Vallini V, Ortori S, Boraschi P, et al. Staging of pelvic lymph nodes in patients with prostate cancer: Usefulness of multiple b value SE-EPI diffusion-weighted imaging on a 3.0 T MR system. Eur J Radiol Open. 20163:16–21.

36. Linderman JJ, Riggs T, Pande M, et al. Characterizing the dynamics of CD4+ T cell priming within a lymph node. J Immunol. 2010;1846):2873–85.

37. Drake CG, Jaffee E, Pardoll DM. Mechanisms of immune evasion by tumors. Adv Immunol. 200690:51–81.

38. Halle S, Halle O, Forster R. Mechanisms and Dynamics of T Cell-Mediated Cytotoxicity In Vivo. Trends Immunol. 2017;386):432–43.

39. Beck RJ, Slagter M, Beltman JB. Contact-Dependent Killing by Cytotoxic T Lymphocytes Is Insufficient for EL4 Tumor Regression In Vivo. Cancer Res. 2019;7913):3406–16.

40. Del Monte U. Does the cell number 10(9) still really fit one gram of tumor tissue? Cell Cycle. 2009;83):505–6.

41. Moreno CC, Mittal PK, Sullivan PS, et al. Colorectal Cancer Initial Diagnosis: Screening Colonoscopy, Diagnostic Colonoscopy, or Emergent Surgery, and Tumor Stage and Size at Initial Presentation. Clin Colorectal Cancer. 2016;151):67–73.

42. Zastrow S, Phuong A, von Bar I, et al. Primary tumor size in renal cell cancer in relation to the occurrence of synchronous metastatic disease. Urol Int. 2014;924):462–7.

43. Ball DL, Fisher RJ, Burmeister BH, et al. The complex relationship between lung tumor volume and survival in patients with non-small cell lung cancer treated by definitive radiotherapy: a prospective, observational prognostic factor study of the Trans-Tasman Radiation Oncology Group (TROG 99.05). Radiother Oncol. 2013;1063):305–11.

44. Vasaturo A, Halilovic A, Bol KF, et al. T-cell Landscape in a Primary Melanoma Predicts the Survival of Patients with Metastatic Disease after Their Treatment with Dendritic Cell Vaccines. Cancer Res. 2016;7612):3496–506.

45. Ahnert K, Mulansky M. Odeint – Solving Ordinary Differential Equations in C++. AIP Conference Proceedings. 2011;1389.

46. Scheffer M, Carpenter SR, Lenton TM, et al. Anticipating critical transitions. Science. 2012;3386105):344–8.

47. Scheffer M, Bascompte J, Brock WA, et al. Early-warning signals for critical transitions. Nature. 2009;4617260):53–9.

48. Eisenhauer EA, Therasse P, Bogaerts J, et al. New response evaluation criteria in solid tumours: revised RECIST guideline (version 1.1). Eur J Cancer. 2009;452):228–47.

49. Champiat S, Dercle L, Ammari S, et al. Hyperprogressive Disease Is a New Pattern of Progression in Cancer Patients Treated by Anti-PD-1/PD-L1. Clin Cancer Res. 2017;238):1920–8.

50. Lesterhuis WJ, Bosco A, Millward MJ, et al. Dynamic versus static biomarkers in cancer immune checkpoint blockade: unravelling complexity. Nat Rev Drug Discov. 2017;164):264–72.

51. Iwai Y, Ishida M, Tanaka Y, et al. Involvement of PD-L1 on tumor cells in the escape from host immune system and tumor immunotherapy by PD-L1 blockade. Proc Natl Acad Sci U S A. 2002;9919):12293–7.

52. Yang B, Li M, Tang W, et al. Dynamic network biomarker indicates pulmonary metastasis at the tipping point of hepatocellular carcinoma. Nat Commun. 2018;91):678.

53. Liu R, Wang J, Ukai M, et al. Hunt for the tipping point during endocrine resistance process in breast cancer by dynamic network biomarkers. J Mol Cell Biol. 2019;118):649–64.

54. Sunshine J, Taube JM. PD-1/PD-L1 inhibitors. Curr Opin Pharmacol. 201523:32–8.

55. Thomas A, Routh ED, Pullikuth A, et al. Tumor mutational burden is a determinant of immune-mediated survival in breast cancer. Oncoimmunology. 2018;710):e1490854.

56. Balar AV, Galsky MD, Rosenberg JE, et al. Atezolizumab as first-line treatment in cisplatin-ineligible patients with locally advanced and metastatic urothelial carcinoma: a single-arm, multicentre, phase 2 trial. Lancet. 2017;38910064):67–76.

57. Blank CU, Haanen JB, Ribas A, et al. CANCER IMMUNOLOGY. The “cancer immunogram”. Science. 2016;3526286):658–60.

58. Galon J, Pages F, Marincola FM, et al. The immune score as a new possible approach for the classification of cancer. J Transl Med. 201210:1.

