## Supplementary Information for "A tipping point in cancer-immune dynamics leads to divergent immunotherapy responses and hampers biomarker discovery"

### 1 Supplementary Figures

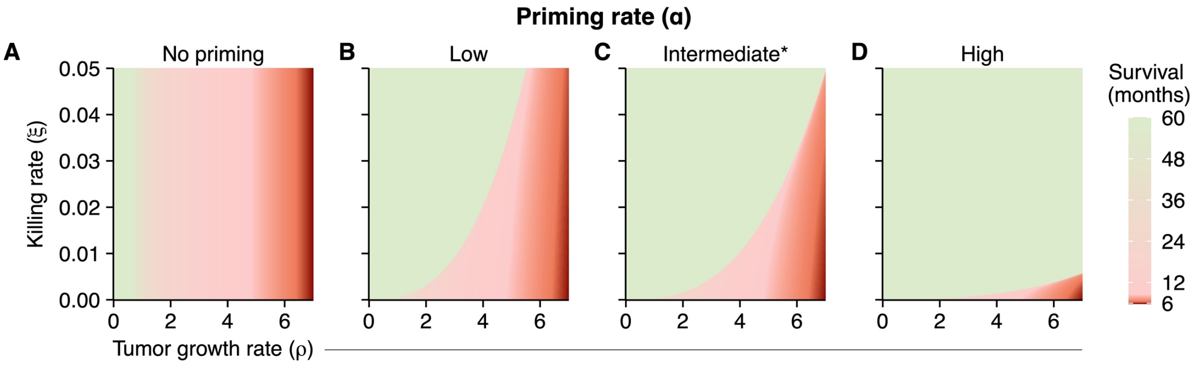


***Supplementary Figure 1: A tumor's priming rate affects the location of a tipping point.***
***(A)*** *In the absence of T cell priming, survival is only determined by the tumor growth rate. Logically, a tipping point cannot be present. Priming rate $\alpha$ = 0.* ***(B-D)*** *Higher T cell priming rates lead to increased availability of cytotoxic T cells. As a result, despite a similar killing rate, the augmented T cell pool can clear tumors with a higher priming rate more easily. These findings are visible as a shifting tipping point in the phase diagrams. As stated in the Methods, a priming rate of 0.0025 is mechanistically plausible and, therefore, selected as the default priming rate (indicated with a *). Parameter values for low and high priming rates are $\alpha$ = 0.00125 and $\alpha$ = 0.025, respectively.*

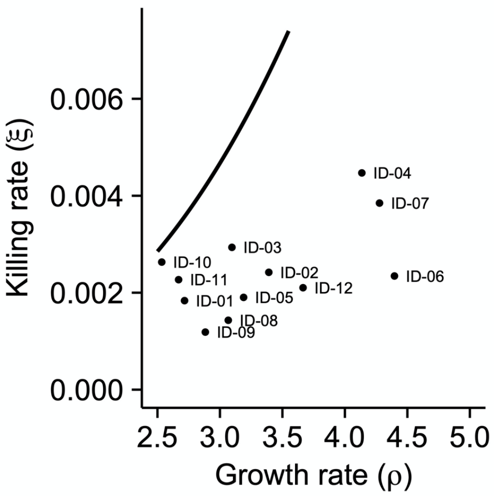


***Supplementary Figure 2: Tumor-immune dynamics determine the clinical outcome of patients in close proximity to a tipping point.***

### 2 Supplementary Tables

*Supplementary Table 1: Simulation parameters of Figure 1*

| Panel | Simulation parameters |
|---|---|
| Overall | $\rho$ = 1 |
| B | $\xi$ = 0.005 |
| C | $\xi$ = 0.00025 |
| D | $\xi$ = 0.0005 |

*Supplementary Table 2: Simulation parameters of Figure 2*

| Panel | Simulation parameters |
|---|---|
| A | Main: $\rho$ = range from 0 to 7, $\xi$ = 0.005<br><br>Inset 1: $\rho$ = 2, $\xi$ = 0.005<br><br>Inset 2: $\rho$ = 5.5, $\xi$ = 0.005 |
| B | Main: $\rho$ = 6, $\xi$ = range from 0 to 0.005<br><br>Inset 1: $\rho$ = 6, $\xi$ = 0.005<br><br>Inset 2: $\rho$ = 6, $\xi$ = 0.035 |
| C | $\rho$ = range from 0 to 7<br>$\xi$ = range from 0 to 0.05 |

*Supplementary Table 3: Simulation parameters of Figure 3*

| Panel | Simulation parameters |
|---|---|
| B/C | Main: $\rho$ = 2, $\xi$ = 0.001<br><br>Variation in treatment effect and treatment duration are indicated on the x-axes of the figures. |
| D | Baseline values for the T cell killing rate were fixed at $\xi$ = 0.0025.<br>Baseline values for the tumor growth rate ($\rho$) were sampled from a normal distribution: $\rho \sim N(2.5, 1)$. We included only patients (n = 20) with clinically evident tumors. |
| E | Baseline values for the tumor growth rate were fixed at $\rho$ = 2.5.<br>Baseline values for the T cell killing rate ($\xi$) were sampled from a uniform distribution: $\xi \sim U$ (0, 0.005). We included only patients (n = 20) with clinically evident tumors. |

*Supplementary Table 4: Simulation parameter of Figure 4*

| Panel | Simulation parameters |
|---|---|
| Overall | Treatment effect = $\xi$ * 12.5 |
| A | $\rho$ = 2, $\xi$ = 0.001, stochasticity tumor growth rate = 0.3 |
| B | $\rho$ = 2, $\xi$ = 0.001, stochasticity T cell killing rate = 0.3 |
| C | Baseline values were sampled from two normal distributions:<br>• $\rho \sim N(2.5, 1)$<br>• $\xi \sim N(0.0025, 0.001)$<br>We select patients (n = 12) with clinically evident tumors and rejected all patients in which tumors did not exceed the diagnosis threshold.<br><br>Stochasticity tumor growth rate = 0.3, stochasticity T cell killing rate = 0.3 |
| D | $\rho$ = range from 0 to 7, $\xi$ = 0.005, stochasticity tumor growth rate = 0.3 |
| E | $\rho$ = 6, $\xi$ = range from 0 to 0.05, stochasticity T cell killing rate = 0.3 |

*Supplementary Table 5: Simulation parameters of Figure 5*

| Panel | Simulation parameters |
|---|---|
| Overall | Baseline values sampled from two uniform distributions:<br>• $\rho \sim U$ (4, 5) |

| | |
|---|---|
| | • ξ ~ *U* (0.015, 0.025)<br><br>Simulations where the tumor did not become clinically apparent (i.e., did not reach a size of 65 * $10^8$ tumor cells) were not included in the analysis. |
| A | Treatment effect = ξ * 4 |
| B | Treatment effect = ξ * 4<br>Stochasticity in tumor growth rate = 0.05<br>Stochasticity in T cell killing rate = 0.05 |

*Supplementary Table 6: Baseline characteristics of retrospective validation cohort.*

| | **Overall (N=58)** |
|---|---|
| **Gender** | |
| Female | 21 (36.2%) |
| Male | 37 (63.8%) |
| **Age (years)** | |
| Median [Min, Max] | 51.0 [19.0, 76.0] |
| **Breslow thickness (mm)** | |
| Median [Min, Max] | 2.65 [0.7, 13.0] |
| **M stage at inclusion** | |
| M1a | 13 (22.4%)* |
| M1b | 14 (24.1%) |
| M1c | 31 (53.4%) |
| **LDH (U/L)** | |
| Median [Min, Max] | 388 [228, 1830] |
| **Time to M stage (months)** | |
| Median [Min, Max] | 29.3 [0, 137] |
| **Overall Survival (months)** | |
| Median [Min, Max] | 8.92 [1.15, 130] |

* Includes one irresectable stage III melanoma patient.

*Supplementary Table 7: Cox proportional hazard models on validation cohort.*

| **Model** | **N** | **HR*** | **95% CI** | **Wald statistic** | **Likelihood ratio test** |
|---|---|---|---|---|---|
| **LDH** | 58 | 6.92 | (2.93 - 16.31) | $p$ = 1.01e$^{-5}$ | $p$ = 4e$^{-5}$ |
| **I/P ratio** | 58 | 0.64 | (0.53 - 0.77) | $p$ = 2.07e$^{-6}$ | $p$ = 3.7e$^{-7}$ |
| **LDH + I/P ratio** | 58 | | | | $p$ = 9.3e$^{-10}$ |
| LDH | | 7.80 | (2.98 - 20.37) | $p$ = 2.8e$^{-5}$ | |
| I/P ratio | | 0.65 | (0.55 - 0.78) | $p$ = 4.4e$^{-6}$ | |

* Before analysis, all predictors were log-transformed.
